# Interventions for burnout and well-being in homelessness staff: a systematic scoping review

**DOI:** 10.1101/2024.08.21.24312389

**Authors:** L Ng, E Adams, D Henderson, E Donaghy, SW Mercer

## Abstract

**Background:** Homelessness staff often experience high job demands, limited resources, and significant emotional strains; with high levels of burnout, stress, and trauma being common within the workforce. Despite growing recognition of these issues, limited information exists regarding interventions to address this.

**Aim:** To conduct a systematic scoping review of interventions aimed at improving well-being and reducing burnout among homelessness staff.

**Methods:** All eligible studies needed to include an intervention addressing burnout and/or well-being in homelessness staff, published in English with primary data. Evidence sources were left open with no data restrictions. Following a registered protocol (available at osf.io/jp5yx), a systematic search of five electronic databases (Medline, APA PsychInfo, Global Health, ASSIA, CINAHL) and Google Scholar was conducted. Studies were double-screened for inclusion. Methodological quality was assessed using the Mixed Methods Appraisal Tool.

**Results:** Out of 5,775 screened studies, six met the inclusion criteria: two peer-reviewed and four non-peer-reviewed publications. No studies were retrieved from Google Scholar. The included studies comprised four quantitative non-randomised designs, one randomised controlled trial, and one mixed-methods study, with four being complex interventions. Three were therapy-based, two included supervision, and two comprised one-time educational sessions. Most were conducted in the United States (n=4), with two in the United Kingdom. The total pooled sample was 347 participants, though four studies were missing demographic data (age and gender). The studies used heterogenous measures and outcomes. Limitations included restrictions to English-only publications, potential gaps in capturing well-being measures, and a limited grey literature scope.

**Conclusion:** There is a lack of research on well-being and burnout interventions in frontline homelessness staff. Identified studies were generally of low quality with a heterogeneity of measures and outcomes used to assess well-being and burnout, limiting generalisability of findings. More robust study designs, along with standardised measures and outcomes, are needed going forwards.

## Introduction

Homelessness is increasing within Europe and the United Kingdom (UK), with increasing demands placed on staff working in homelessness services.^1,2^ With the rising numbers of people experiencing homelessness (PEH), there has been growing recognition that the well-being of homelessness staff is crucially important in providing high-quality care for PEH.^3–5^ Although staff working in the homelessness sector often find the work rewarding, it is nevertheless acknowledged to be challenging, with the workforce facing high levels of staff turnover, stress, burnout, and secondary trauma.^3,6,7^

Homelessness staff often endure high job demands, limited resources, in addition to emotional health strains.^7^ PEH often have complex histories, intertwined with previous or current exposure to trauma, abuse, violence, substance misuse and mental-health concerns.^8^ Homelessness staff are at risk of experiencing vicarious trauma or secondary traumatic stress as a result of this exposure to trauma,^9^ in addition to their own personal histories as well.^10^ A recent study highlighted that adverse childhood experiences among homelessness staff are higher compare to the general population, which may increase susceptibility to and burnout if not appropriately supported.^10^ Moreover, broader systemic issues, such as resource disparity, insufficient funding, low wages and organisational silos between professional groups caring for PEH, can further hinder the ability of practitioners to provide appropriate biopsychosocial care for PEH.^11,12^

While factors contributing to the mental health of homelessness staff are being increasingly researched, little remains known regarding the interventions that have been evaluated to address this. Pressing calls to explore this gap have been made.^13,14^

To move the field forward, an understanding of the existing research on interventions is needed. Therefore, the objective of this systematic scoping review is to map and identify well-being and burnout interventions implemented for homelessness staff.

## Methods

A systematic scoping review approach was adopted to answer the wider research question, namely to identify the extent and nature of existing research and to ascertain the methodologies used to conduct these interventions.

### Study design

This scoping review was conducted in accordance to the Joanna Briggs Institute methodology for scoping reviews,^15^ based on Arksey and O’Malley^16^ and Levac et al’s^17^ framework. The review is reported using the PRISMA extension for Scoping Reviews (PRISMA-ScR) flow diagram.^18^ The review protocol was registered on Open Science Framework (OSF) in May 2023 (S1 File).^19^

### Research questions

This scoping review addressed the following questions:

1. What interventions have been implemented in the homelessness sector to address staff well-being and burnout?

a. In what settings and context were these interventions carried out?
b. What measurement tools and outcomes were used to evaluate well-being and burnout in these studies?
c. How did the interventions change practice?

### Eligibility criteria

For the purposes of this review, well-being included any intervention addressing stress, burnout, job satisfaction, compassion fatigue, secondary traumatic stress, vicarious trauma, post-traumatic stress and well-being itself. These aspects have previously been identified as part of the emotional pressures faced among homelessness staff.^13,14^

The inclusion criteria followed the Population, Concept, and Context criteria (see Table 1). Studies were selected if they met the following three criteria: (1) the intervention specifically addressed burnout and well-being in homelessness staff and/or trainees; (2) full-text was available in the English language; and (3) the evaluation contained primary data. Evidence sources were left open, with no date restrictions.

**Table 1.** Study criteria (Population, Concept, Context and Evidence sources)

|  | **Inclusion criteria** | **Exclusion criteria** |
| --- | --- | --- |
| **Participants** | Any support staff and trainees working in direct contact with PEH. | No direct contact with PEH |
| **Concept** | Any interventions that address burnout and well-being in homelessness workers. | Interventions not addressing burnout or well-being |
| **Context** | Any organisation supporting PEH | Organisations that do not work with PEH |
| **Evidence sources** | Any research assessing primary data in the English language<br>No restrictions with source type or publication date | Non-English study, which has not been translated<br>No primary data |

### Information sources and strategy

An initial search of Medline, PsychInfo, Global Health, ASSIA and CINAHL was undertaken to identify articles relating to the review. In addition, recommended search strategies from a related systematic review and scoping review were used to supplement the initial scoping searches.^13,14^ An academic librarian was subsequently consulted to help refine the search terms and databases.

The final search strategy included five electronic databases: Medline, PsychInfo, Global Health, ASSIA, CINAHL. The search strategy was conducted on August 28^th^, 2023 by LN in English, due to language limitations of reviewers, and adapted to each database, with no date limitations. To identify any additional studies, Google Scholar was searched using the following terms: (“burnout”) and (“homeless”) and (“staff”) and (“intervention”). The first 300 articles in the Google Scholar search to appear were included in the screening. References of the final included sources were also screened for supplementary articles; however, none were identified. An example search string from Medline and PsychInfo is shown in Table 2.

**Table 2.** Example search string from PsychoInfo and Medline databases on the OVID platform.

| Database | Search string |
| --- | --- |
| **An example search of APA PsychInfo and OVID Medline** | APA PsycInfo <1806 to May Week 3 2023><br>Ovid MEDLINE(R) ALL <1946 to May 22, 2023> |
|  | 1 ((Burnout or stress* or "emotional* exhaust*" or workload* or "vicarious trauma*" or "compassion fatigue" or "secondary trauma*" or PTSD or "post-trauma* stress" or "posttrauma* stress" or depression or "mental health" or "well-being" or wellbeing or "job satisfaction" or "job dissatisfaction" or resilience or coping or "self-efficacy") adj4 (work* or professional* or employe* or staff or personnel* or manager*)).mp. [mp=ti, ab, hw, tc, id, ot, tm, mf, bt, nm, fx, kf, ox, px, rx, an, ui, sy, ux, mx] 214144 |
|  | 2 (homeless* or houseless* or "street dwell*" or "shelter dwell*" or "street youth*" or "street people" or "street child*" or "street person*" or unhoused or unsheltered or "rough sleep*" or "sleep* rough" or runaway* or "supported housing" or "fixed abode" or "ill-housed" or vagrant* or "people living on the street*" or "sofa surf*" or shelter*).mp. [mp=ti, ab, hw, tc, id, ot, tm, mf, bt, nm, fx, kf, ox, px, rx, an, ui, sy, ux, mx] 54999 |
|  | 3 (intervention* or program* or education or training or workshop* or course* or curriculum or approach* or service* or "random* control* trial*" or rct* or "experimental design*").mp. [mp=ti, ab, hw, tc, id, ot, tm, mf, bt, nm, fx, kf, ox, px, rx, an, ui, sy, ux, mx] 9197235 |
|  | 4 1 and 2 and 3 839 |

### Study selection

Our search initially yielded 8,447 articles, in addition to 300 articles retrieved from Google Scholar. SR-Accelerator was used to remove any initial duplicates, with further duplicates removed by Covidence or manually by a reviewer. Search results were uploaded onto Covidence. Inclusion and exclusion criteria were completed by the primary reviewer (LN). Four reviewers (LN, EA, DA, ED) completed Title and Abstract screening independently and two reviewers (LN and EA) completed Full Text review and Data Extraction independently. Two reviewers, including the primary reviewer (LN), independently assessed the papers and identified if they met the inclusion criteria. Where there were discrepancies in study selection, a third and fourth reviewer (SM and EA) adjudicated on the final decision. Fig 1 summarises the screening process and reasons for exclusion.

**Figure 1.**
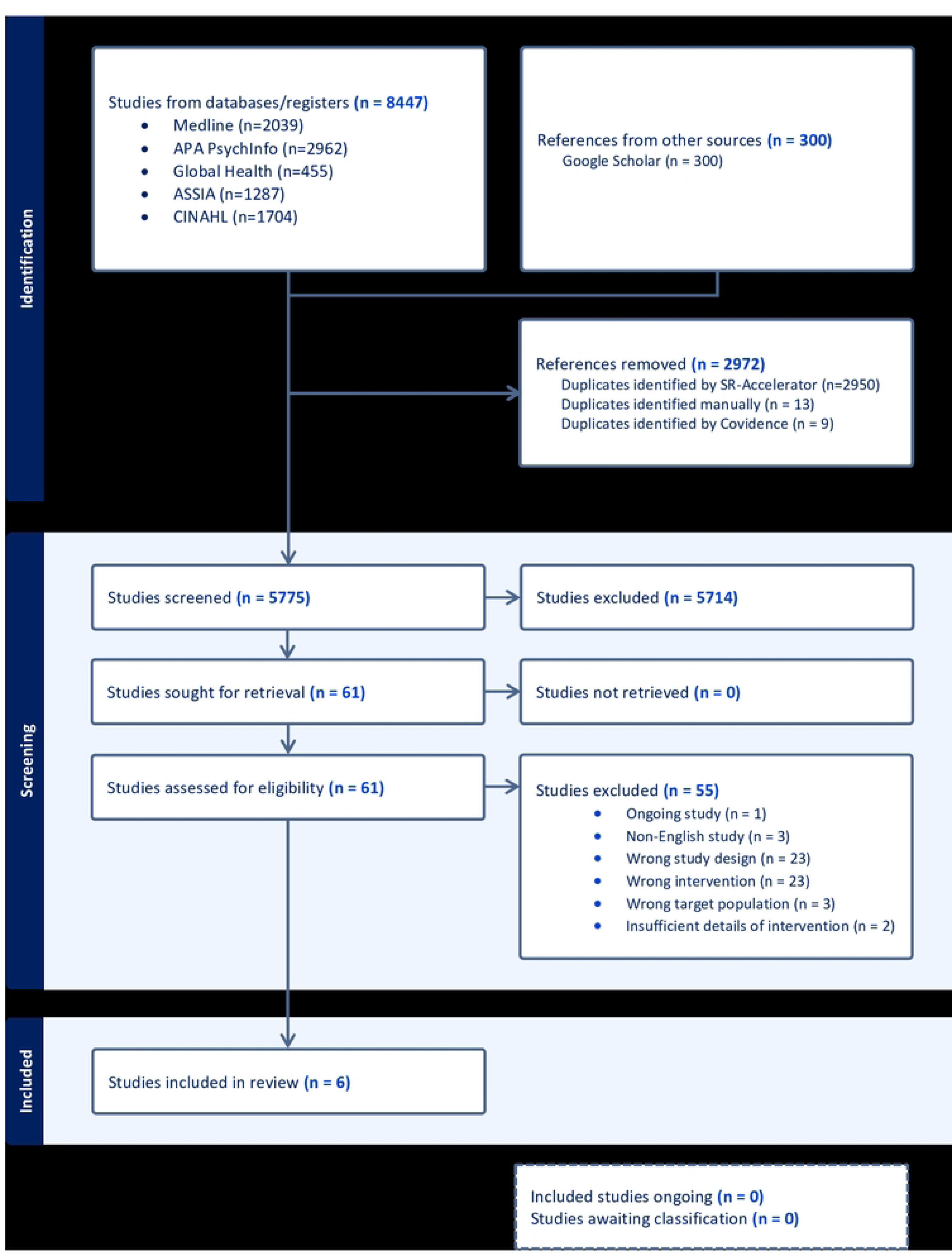
PRISMSA flowchart of scoping review.

### Quality assessment

Quality assessment of the final included studies (n=6) was carried out by the primary reviewer (LN), using the Mixed Methods Appraisal Tool (MMAT).^21^ While papers were not excluded based on poor quality, the tool was used to the ascertain if the methodologies were appropriately carried out and to make an informed judgment of its findings.

## Results

After de-duplication, a total of 5,775 studies were screened. After screening, 61 studies were reviewed at full-text. Of these, 6 were eligible for the review.

### Study characteristics

The study characteristics are summarised in Table 3. Most studies were not published in peer-reviewed journals (n=4),^21–24^ with only two undergoing peer-review (n=2).^25,26^ Among the non-peer-reviewed studies, three were dissertations published in an online database^21,23,24^ and one was an unpublished manuscript from an institutional repository.^22^ No included studies were retrieved from Google Scholar. Nearly all were published in the last fifteen years (n=5).^21–23,25,26^ Most were conducted in the United States (US) (n=4)^21,23–25^, with the remaining two conducted in United Kingdom (UK).^22,26^ Information on participant demographics, namely age and/or gender, were missing in four of the six studies.^22,23,25,26^ From the available data, most participants were female. The settings where the studies took place included specialist homelessness organisations (n=3)^21,22,26^, a domestic violence (DV) shelter (n=1),^24^ a medical home for veterans experiencing homelessness (n=1),^25^ and a community healthcare organisation for underserved populations, including PEH (n=1).^23^ Quantitative pre-experimental design was the most common study design used (n=4).^21,22,25,26^ Only one study used a randomised control trial (RCT) design^24^ and one used a mixed-methods non-experimental design.^23^

**Table 3.** Characteristics of included publications (n=6)

| Author | Publication characteristics<br>1. Peer-review status<br>2. Type<br>3. Publication source (Name) | Setting<br>1. Service delivery setting<br>2. Type of service users<br>3. Country | Participant sample<br>1. Age (mean ± SD)<br>2. Gender (participant number)<br>3. Job role(s)<br>4. Total number of participants (dropouts) | Methods<br>1. Study design<br>2. Data collection |
| --- | --- | --- | --- | --- |
| **Demasi (2023)** | 1. Non-peer-reviewed<br>2. Dissertation for Doctor of Nursing Practice<br>3. Research Database (ProQuest) | 1. A faith-based, non-profit organisation working with PEH<br>2. PEH<br>3. United States | 1. 54 years (11)<br>2. Male (4) and Female (12)<br>3. Volunteers<br>4. 23 total (7 dropouts) | 1. Quantitative non-randomised<br>2. Cross-sectional |
| **Jeffrey (1999)** | 1. Non-peer-reviewed<br>2. Dissertation for Doctor of Philosophy in Psychology<br>3. Research Database (Dissertation Abstracts International: Section B: The Sciences and Engineering) | 1. Various shelter workers working with victims of domestic violence (DV)<br>2. DV<br>3. United States | 1. 36 years (11)<br>2. 93% female<br>3. A range of DV shelters workers (eg. counsellors, program directors, administration)<br>4. 267 total (195 dropouts) | 1. Quantitative randomised control trial<br>2. Cross-sectional |
| **Maguire et al. (2017)** | 1. Non-peer-reviewed<br>2. Unpublished manuscript<br>3. Institutional Repository (University of Southampton Institutional Repository) | 1. 17 homeless organisations in London<br>2. PEH ('rough sleepers')<br>3. United Kingdom | 1. Not stated<br>2. Male (13) and Female (17)<br>3. Homelessness staff<br>4. 30 staff (15 dropouts) | 1. Quantitative non-randomised<br>2. Longitudinal |
| **Moore et al. (2019)** | 1. Peer-reviewed<br>2. Research article<br>3. Academic Journal (The Clinical Teacher) | 1. A patient-centred medical home providing healthcare, housing and social resources to veterans experiencing homelessness<br>2. Veterans experiencing homelessness<br>3. United States | 1. Not stated<br>2. Not stated<br>3. Healthcare trainees (doctors, nurses, pharmacists and psychologists)<br>4. 15 total (7 dropouts from post implementation survey & 3 dropouts from value assessment tools) | 1. Quantitative non-randomised<br>2. Cross-sectional |
| **Munyoki (2022)** | 1. Non-peer-reviewed<br>2. Dissertation for Doctor of Nursing Practice<br>3. Research Database (ProQuest) | 1. A community organisation healthcare practice for low income and medically underserved populations, including PEH<br>2. Vulnerable underserved populations, including PEH in a homelessness shelter site<br>3. United States | 1. Mean not stated, 60% participants were in 40-49 years age group.<br>2. Not stated<br>3. A range of staff (eg. social workers and nurse practitioners)<br>4. 8 total (4 dropouts) | 1. Mixed-methods non-experimental design<br>2. Not applicable |
| **Reeve et al. (2021)** | 1. Peer-reviewed<br>2. Research article<br>3. Academic Journal (Clinical Psychology & Psychotherapy) | 1. A homelessness organization in East Midlands<br>2. PEH<br>3. United Kingdom | 1. Not stated<br>2. Female (4), no male<br>3. 2 support development workers and 2 assistant managers<br>4. 4 total (no dropouts) | 1. Quantitative non-randomised<br>2. Longitudinal |

### Interventions

All interventions varied in nature, with their respective components detailed in Table 4. Five were complex interventions,^21,22,24–26^ defined as interventions consisting of several interacting components and measuring multiple outcomes.^27^ Three interventions involved therapy components, namely cognitive behavioural therapy (CBT),^22^ mindfulness,^23^ and acceptance and commitment therapy.^26^ Two of the interventions comprised of educational sessions, one of which involved a session on self-care^21^ and the other presenting a well-being toolkit.^25^ Two of the sessions also incorporated elements of supervision in the intervention, namely feedback on secondary traumatic stress for homelessness staff^24^ and psychologist supervision for CBT training.^22^ Four of the six interventions completed mainly in-person.^21,22,25,26^ One intervention involved delivering a mindfulness intervention through an online platform^23^ and another intervention used an anonymous postal feedback survey for homelessness staff on secondary traumatic stress symptoms.^24^ Most interventions were evaluated over one to three months.^21,23,24,26^ The longest evaluation period was over an academic year, estimated to be approximately 8-10 months, although the exact duration in months was not specified in the study.^25^

**Table 4.** Intervention components and measures.

| Author | Intervention name | Complex Intervention (Yes/No) | Components | Duration (Months) | Outcomes and Measures<br>Outcomes: Measures |
| --- | --- | --- | --- | --- | --- |
| **Demasi (2023)** | An in-person educational session on self-care activities | Yes | ● 50-minute educational session on compassion fatigue and self-care<br>● Self-care tool box (eg. mindfulness leaflets) – accessed as needed | 1 month | ● *Compassion fatigue, burnout and secondary traumatic stress:* Professional Quality of Life V Scale (ProQOL) |
| **Jeffrey (1999)** | Feedback intervention on Secondary Traumatic Stress (STS) | Yes | ● Providing feedback regarding STS levels of staff<br>● Each individual staff member was assigned either control group (CG), Feedback Only Group (FG), or Feedback Intervention Group (FIG).<br>● CG received no feedback. FG and FIG received individual and director reports of STS. FIG additionally received a list of suggestions to address STS.<br>● Individual feedback involved statements provided to workers about their performance compared to others.<br>● Director feedback, provided only to shelter directors, included information of the shelter's performance compared to other DV shelters and information on the measures. | 2 months | ● *Post-traumatic stress disorder (PTSD):* Modified PTSD Symptom Scale<sup>b</sup><br>● *Impact of PTSD:* Impact of Events Scale (IES)<sup>b</sup><br>● *Beliefs and schemas about self and others due to vicarious trauma:* Traumatic Stress Institute Belief Scale (TSI) (Revision L)<br>● *Coping skills:* Coping Strategies Inventory (CSI)<br>● *Implementing coping skills to help with PTSD:* Assessment of Coping with Traumatic Stress (ACTS) |
| **Maguire et al. (2017)** | Cognitive-behavioural therapy (CBT) training and supervision package | Yes | ● Designed and led by clinical psychologist<br>● 4-day CBT skills workshops to increase workers skills in CBT, and reduce burnout and negative attitudes<br>● Psychologist-led supervision (in groups of three staff) in CBT training every 2 weeks for 6 months | 6 months | ● *Burnout:* Maslach Burnout Inventory (MBI)<br>● *Negative beliefs about clients:* Staff Attitudes and Beliefs Questionnaire – 42 (SAB42)<sup>b</sup> |
| **Moore et al. (2019)** | A well-being toolkit intervention | Yes | ● Well-being toolkit with evidence-based tools, led by national expert (eg. well-being practices, booklets)<br>● A 'Wellness Room' (quiet space)<br>● Daily gratitude practice in morning huddles<br>● Half-day workshop on toolkit use | 8-10 months<sup>a</sup> | ● *Burnout:* MBI (only one scale used to avoid survey fatigue but scale not specified)<br>● *Stress:* Cohen Perceived Stress Scale<br>● *Resilience:* Connor-Davidson Resilience Scale<br>● *Mindfulness:* Five-Factor Mindfulness Scale (only two of the five subscales to avoid survey<br>fatigue: 'Nonreactivity to Inner Experience' and 'Observing')<br>● *Assessing the value of each tool*: Bespoke Likert scales<sup>b</sup> |
| **Munyoki (2022)** | An abbreviated version of the 8-week Mindfulness-based stress reduction (MBSR) program | No | ● Presentation of MBSR program<br>● Mindfulness modules on online platform (at least 15-40 minutes, three times a week) | 1 month | ● *Assessing the intervention*: Bespoke quantitative survey questions (Likert scales, multiple choice, ranking questions)<sup>b</sup><br>● *Barriers and future recommendations for the intervention*: Free-text boxes with qualitative data<sup>b</sup> |
| **Reeve et al. (2021)** | Acceptance and commitment therapy (ACT) | Yes | ● Project advertised in team meetings and personal communication<br>● ACT-intervention was split into three modules reflecting the three aspects of ACT triflex ('being open', 'noticing' and 'being active') and conducted as one-to-one workshop-style sessions | 2-3 months | ● *Burnout*: Oldenburg Burnout Inventory (OLBI)<br>● *Well-being*: Personal well-being index (PWI)<br>● *Psychological Flexibility*: Comprehensive assessment of Acceptance and Commitment Therapy processes (CompACT)<br>● *Idiographic personal value*: An untested single-item measure asking participants to identify one meaningful value from home and one from work<sup>b</sup> |

<sup>a</sup>The paper states the intervention took place over an academic year but no specific timeframe was indicated.
<sup>b</sup>These are non-validated measures.

### Outcomes and measures

The outcomes and measures used to assess well-being in homelessness staff are listed in Table 4.

With regards to outcomes, four studies assessed burnout.^21,22,25,26^ Two studies assessed staff beliefs on self and/or others, including service users.^22,24^ Two studies evaluated general well-being of staff^21,26^ and two studies evaluated coping abilities.^24,26^ Two interventions used bespoke questions assessing the interventions themselves.^23,25^ One study used stress and resilience as an outcome measure^25^ and another study used PTSD symptoms as an outcome measure.^24^

Nearly all measures used to evaluate well-being varied in the six studies. Only two studies used the same measurement tool, the Maslach Burnout Inventory (MBI).^22,25^ Only one of the MBI scales were used in Moore et al.’s study.^25^ Moreover, almost all studies incorporated non-validated measures in their evaluation.^22–26^

### Quality appraisal

Nearly all studies had methodological limitations, mainly owing to small sample sizes, high drop-out rates, insufficient details on the study’s recruitment and methodology, use of non-validated measures, and lack of accounting for confounding variables (S1 Table). Based on the MMAT criteria,^20^ four of the six studies scored between 0-20% in methodological quality,^22–25^ one study scored 40%,^21^ and one met all of the appraisal criteria.^26^ Although one RCT evaluation was included,^24^ the quality was poor and lacked rigor to draw conclusions from its findings. No power calculations were conducted in any of the studies. However, Reeve et al’s study^26^ identified that a minimum of three participants were required for establishing an effect in single-case experimental design research, as used in their study, and highlighted that their study met this respective criterion.

### Key findings and recommendations

The study’s key findings and future recommendations are shown in Table 5. The results of three studies reached statistical significance.^21,22,26^ Two interventions demonstrated statistically significant improvements in burnout among homelessness staff following an in-person educational session on self-care,^21^ and following CBT training and supervision.^22^ The Acceptance and Commitment Therapy intervention illustrated statistically significant increased psychological flexibility in half of the participants.^26^

**Table 5.**
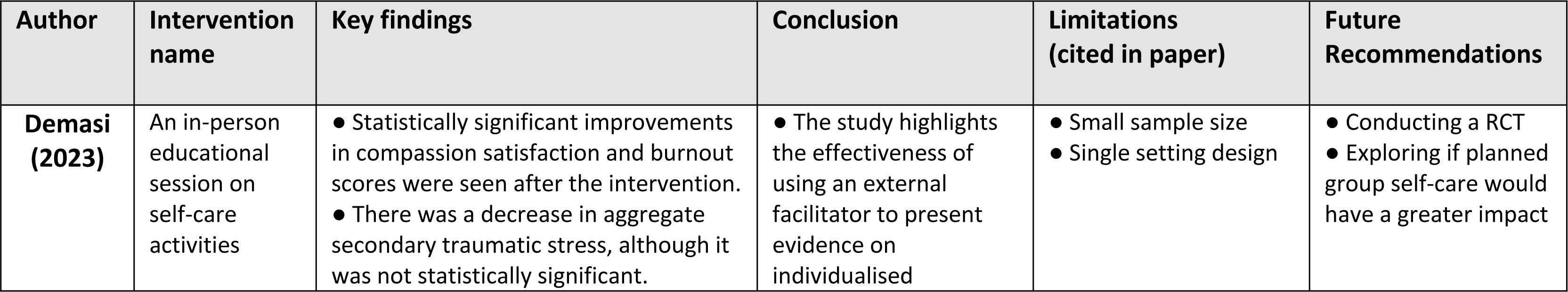
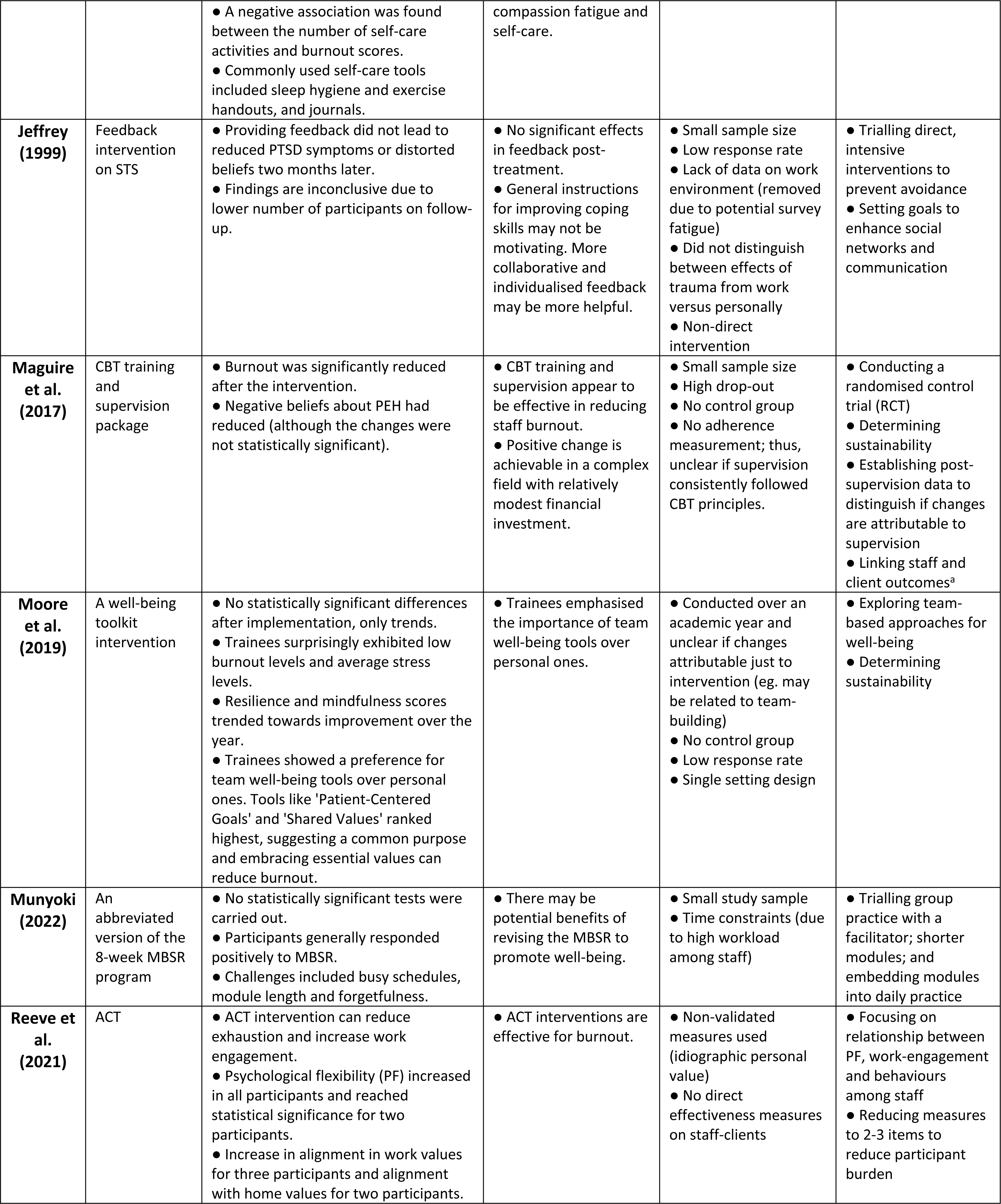
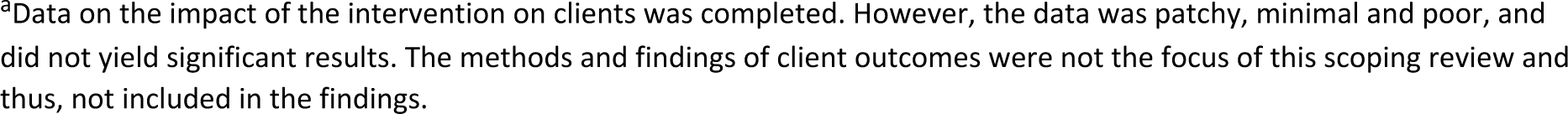
Intervention key findings and implications on practice.

| Author | Intervention name | Key findings | Conclusion | Limitations (cited in paper) | Future Recommendations |
| --- | --- | --- | --- | --- | --- |
| **Demasi (2023)** | An in-person educational session on self-care activities | Statistically significant improvements in compassion satisfaction and burnout scores were seen after the intervention.<br>There was a decrease in aggregate secondary traumatic stress, although it was not statistically significant. | The study highlights the effectiveness of using an external facilitator to present evidence on individualised | Small sample size<br>Single setting design | Conducting a RCT<br>Exploring if planned group self-care would have a greater impact |
|  |  | ● A negative association was found between the number of self-care activities and burnout scores.<br>● Commonly used self-care tools included sleep hygiene and exercise handouts, and journals. | compassion fatigue and self-care. |  |  |
| **Jeffrey (1999)** | Feedback intervention on STS | ● Providing feedback did not lead to reduced PTSD symptoms or distorted beliefs two months later.<br>● Findings are inconclusive due to lower number of participants on follow-up. | ● No significant effects in feedback post-treatment.<br>● General instructions for improving coping skills may not be motivating. More collaborative and individualised feedback may be more helpful. | ● Small sample size<br>● Low response rate<br>● Lack of data on work environment (removed due to potential survey fatigue)<br>● Did not distinguish between effects of trauma from work versus personally<br>● Non-direct intervention | ● Trialling direct, intensive interventions to prevent avoidance<br>● Setting goals to enhance social networks and communication |
| **Maguire et al. (2017)** | CBT training and supervision package | ● Burnout was significantly reduced after the intervention.<br>● Negative beliefs about PEH had reduced (although the changes were not statistically significant). | ● CBT training and supervision appear to be effective in reducing staff burnout.<br>● Positive change is achievable in a complex field with relatively modest financial investment. | ● Small sample size<br>● High drop-out<br>● No control group<br>● No adherence measurement; thus, unclear if supervision consistently followed CBT principles. | ● Conducting a randomised control trial (RCT)<br>● Determining sustainability<br>● Establishing post-supervision data to distinguish if changes are attributable to supervision<br>● Linking staff and client outcomes<sup>a</sup> |
| **Moore et al. (2019)** | A well-being toolkit intervention | ● No statistically significant differences after implementation, only trends.<br>● Trainees surprisingly exhibited low burnout levels and average stress levels.<br>● Resilience and mindfulness scores trended towards improvement over the year.<br>● Trainees showed a preference for team well-being tools over personal ones. Tools like 'Patient-Centered Goals' and 'Shared Values' ranked highest, suggesting a common purpose and embracing essential values can reduce burnout. | ● Trainees emphasised the importance of team well-being tools over personal ones. | ● Conducted over an academic year and unclear if changes attributable just to intervention (eg. may be related to team-building)<br>● No control group<br>● Low response rate<br>● Single setting design | ● Exploring team-based approaches for well-being<br>● Determining sustainability |
| **Munyoki (2022)** | An abbreviated version of the 8-week MBSR program | ● No statistically significant tests were carried out.<br>● Participants generally responded positively to MBSR.<br>● Challenges included busy schedules, module length and forgetfulness. | ● There may be potential benefits of revising the MBSR to promote well-being. | ● Small study sample<br>● Time constraints (due to high workload among staff) | ● Trialling group practice with a facilitator; shorter modules; and embedding modules into daily practice |
| **Reeve et al. (2021)** | ACT | ● ACT intervention can reduce exhaustion and increase work engagement.<br>● Psychological flexibility (PF) increased in all participants and reached statistical significance for two participants.<br>● Increase in alignment in work values for three participants and alignment with home values for two participants. | ● ACT interventions are effective for burnout. | ● Non-validated measures used (idiographic personal value)<br>● No direct effectiveness measures on staff-clients | ● Focusing on relationship between PF, work-engagement and behaviours among staff<br>● Reducing measures to 2-3 items to reduce participant burden |

<sup>a</sup>Data on the impact of the intervention on clients was completed. However, the data was patchy, minimal and poor, and did not yield significant results. The methods and findings of client outcomes were not the focus of this scoping review and thus, not included in the findings.

In contrast, no statistically significant differences were seen in secondary traumatic stress levels following feedback^24^ and after implementation of the well-being toolkit,^25^ although the latter did demonstrate trends towards improvement in resilience and mindfulness over 8-10 months. A downward trend in secondary traumatic stress scores were seen following the in-person educational session on self-care, however the results were not statistically significant.^21^ Similarly, after CBT training and supervision, there was a reduction in negative beliefs about PEH, but these did not reach statistical significance.^22^

One study did not carry out any statistically significant tests.^23^ However, this was the only study that examined sustainability, rather than outcomes, and highlighted that time and workload were barriers to completing the mindfulness modules.^23^

In nearly all studies, the most common recommendation was exploring the role of group interventions, rather than individual approaches (n=4).^21,23–25^ Other recommendations included conducting randomised controlled trials to isolate the effects of the intervention (n=2),^21,22^ and determining sustainability of the intervention.^22,25^ No adverse events were reported in any studies.

## Discussion

This scoping review evaluated the existing evidence for burnout and well-being interventions for homelessness staff. Of the 5,775 studies screened, a total of six were identified. Only two were published in peer-reviewed research journals. Four studies used quantitative non-randomised designs, one was an RCT, and one used a mixed-methods design. Five consisted of complex interventions, comprising multiple interacting components and targeting multiple outcomes. Three interventions involved therapy components, two comprised one-time educational sessions, and two incorporated elements of supervision in the intervention. Most were conducted in the US, with two completed in the UK. All studies used a wide range of measures and outcomes, with some measures showing statistically significant results. However, applying the MMAT criteria, the study quality was generally poor, owing to small sample sizes; high drop-out rates; poorly characterised participant details, recruitment and methodology; use of non-validated measures, and lack of accounting for confounding variables.

### Strengths and limitations

The strengths of this study include its systematic and in-depth approach to ensure a high-quality study was conducted. Its broad inclusion criteria, search of five databases and Google Scholar, with no time limitations, ensuring a diverse range of papers were assessed. The use of an academic librarian to assist in the search strategy helped reduce the chance of bias in the review. Furthermore, the quality assessment conducted guided the reviewers’ ability to determine whether meaningful conclusions could be drawn.

However, the study was limited to English language publications, potentially missing evidence in other languages. Due to language limitations of reviewers, it was not possible to broaden this further. Moreover, the multidimensional nature of well-being may not have been fully captured in the search strategy, potentially leaving gaps in the literature search. The reviewers attempted to address this by reviewing terms from key papers and recent similar reviews, with regular input from an academic librarian, to ensure a wide and inclusive search scope was upheld. Furthermore, the inclusion of DV shelter workers, while justified by their insecure housing status,^28^ may have introduced some contextual differences. Nonetheless, similar themes to that of homelessness staff have been identified in the literature on DV support workers, including high work demands, burnout, compassion fatigue, and secondary traumatic stress.^29^ Finally, grey literature searches were limited to Google Scholar, which may have omitted some relevant sources. While no studies from Google Scholar were ultimately included in this review, adopting a systematic approach to grey literature searches in future reviews would help ensure that all relevant evidence is captured.^30^

### Comparison with literature

Of the three studies that achieved statistical significance, two were therapeutic interventions, involving relational components facilitated by clinical psychologists,^22,26^ suggesting a potential benefit for clinical psychologist roles in homelessness settings. A recent qualitative study similarly highlighted the value of on-site trainee clinical psychologists in homelessness settings, in terms of providing staff support and promoting psychologically-informed approaches.^31^

Notably, the most common recommendation from studies was to explore group interventions.^21,23–25^ A scoping review on vicarious trauma correspondingly highlighted that group interventions foster group cohesion and support, which helps mitigate secondary traumatic stress symptoms.^32^ Additionally, the ‘Florence Nightingale effect’ suggests that staff who strongly identify with their organization may experience lower burnout and higher job satisfaction when they view client suffering through a lens of organizational commitment, rather than as a traumatic event.^33^ Thus, enhancing organisational identification through group interventions could be a valuable approach for reducing burnout and improving well-being in the homelessness sector.

### Implications for research and practice

All studies employed a wide heterogeneity of measures and outcomes to evaluate well-being, with only two studies using the same measurement tool, and several relying on non-validated tools. The lack of standardisation meant comparability across studies was not possible, and outcome accuracy and reliability were difficult to assess. To improve future research, it is essential to identify and agree upon the most validated measure(s) for assessing well-being and burnout among homelessness staff. A Delphi exercise could be an effective approach to achieving consensus on the most appropriate measures for this group.^34^

Furthermore, most studies were generally low quality, with poorly characterised demographics and methodologies, small sample sizes, and no power calculations, limiting reliability of conclusions. The majority were also quantitatively focused with minimal qualitative insights, leaving the underlying barriers and facilitators of intervention engagement unclear.^35^ Many studies also employed single-group designs with short follow-up periods, making it difficult to assess the full effects and sustainability of interventions. Researchers have underscored appropriate follow-up times are crucial to capture the full impact and sustainability of interventions addressing well-being.^36^ Future research should prioritise robust study designs, with adequate power calculations and follow-up periods, to appropriately capture intervention effects. Incorporating Medical Research Council’s complex intervention guidelines will also enhance study rigor by ensuring interventions are systematically developed, evaluated, and refined. This approach would more effectively capture the complexity of intervention effects and provide more reliable evidence for improving well-being.^27^

## Conclusion

This scoping review shows limited evidence on well-being and burnout interventions for frontline homelessness staff. Studies were generally of low quality with diverse measures used, limiting the ability to draw meaningful conclusions. Robust study designs, such as mixed methods or RCTs, with appropriate power calculations and standardised measures to determine a true effect of an intervention, are needed to guide future interventions on well-being and burnout within the homelessness sector. Incorporating the Medical Research Council guidance on complex interventions will ensure interventions are rigorously developed and evaluated to meet the specific needs of this sector.^27^

## Data Availability

The data in this systematic review are from previously published studies, available through the references listed in the manuscript. These references provide the necessary data for the analyses performed in this review and are accessible as follows: Demasi RR. Helping the Helpers: Self-Care to Impact Compassion Fatigue in Volunteers Working With the Homeless [Internet] [Dissertation]. Carlow University 2023. Available from: https://www.proquest.com/dissertations-theses/helping-helpers-self-care-impact-compassion/docview/2819153475/se-2?accountid=10673 Maguire N, Grellier B, Clayton K. The impact of CBT training and supervision on burnout, confidence and negative beliefs in a staff group working with homeless people. University of Southampton Institutional Repository [Internet]. 2017. Available from: https://eprints.soton.ac.uk/155113/ Munyoki N. Evaluation of a Mindfulness-Based Stress Reduction Program to Reduce Stress, Burnout, and Insomnia for Behavioral Healthcare Staff [Internet] [Dissertation]. University of North Carolina 2022. Available from: https://www.proquest.com/dissertations-theses/evaluation-mindfulness-based-stress-reduction/docview/2671623364/se-2?accountid=10673 Jeffrey AC. Effect of feedback on levels of secondary traumatization of workers at battered women’s shelters across the United States [Internet] [Dissertation]. Virginia Polytechnic Institute and State University 1999. Available from: https://vtechworks.lib.vt.edu/server/api/core/bitstreams/e6832175-bc04-4d0f-989b-0b362e2f2c38/content Moore E, Soh M, Stuber M, Warde C. Well-being for trainees caring for homeless veterans. The Clinical Teacher. 2019 Jul 1116(4). doi: 10.1111/tct.13053 Reeve A, Moghaddam N, Tickle A, Young D. A brief acceptance and commitment intervention for work-related stress and burnout amongst frontline homelessness staff: A single case experimental design series. Clinical Psychology & Psychotherapy. 2021 Mar 2528(5). doi: 10.1002/cpp.2555.

## Acknowledgements

We would like to gratefully acknowledge Rowena Stewart, Academic Support Librarian at University of Edinburgh, for her support and contributions to the literature search.

## Supporting Information

**S1 Table. Results of the quality assessment using the Mixed Methods Appraisal Tool (Hong et al, 2018)**

S1 Table Footnotes: **Abbreviations:** Π = criteria met; Ο = criteria not met, CT = can’t tell due to insufficient information

**S1 File. Well-being and burnout interventions for frontline homelessness staff: A Scoping Review Protocol (registered on OSF:** https://osf.io/jp5yx/)

## References

1. Tunstall R. European Social Policy Network (ESPN): Thematic Report on National strategies to fight homelessness and housing exclusion: United Kingdom [Internet]. Brussels: European Commission; 2019 [cited 2024 Jul 29]. Available from: https://ec.europa.eu/social/BlobServlet?docId=21611&langId=en

2. Foundation Abbe Pierre, FEANTSA. Fifth Overview of Housing Exclusion in Europe 2020 [Internet]. FEANTSA. FEANTSA; 2020 Jul [cited 2024 Jul 29]. Available from: https://www.feantsa.org/public/user/Resources/resources/Rapport_Europe_2020_GB.pdf

3. Manning RM, Greenwood RM. Microsystems of Recovery in Homeless Services: The Influence of Service Provider Values on Service Users’ Recovery Experiences. American Journal of Community Psychology. 2018 Jan 11;61(1-2):88–103. doi:10.1002/ajcp.12215

4. Lemieux-Cumberlege A, Taylor EP. An exploratory study on the factors affecting the mental health and well-being of frontline workers in homeless services. Health & Social Care in the Community. 2019 Mar 12;27(4). doi:10.1111/hsc.12738

5. Gaboardi M, Santinello M, Disperati F, Lenzi M, Vieno A, Loubière S, et al. Working with People Experiencing Homelessness in Europe. Human Service Organizations: Management, Leadership and Governance. 2022 Mar 29;46(4):324–45. doi: 10.1080/23303131.2022.2050330

6. Lemieux-Cumberlege AH, Griffiths H, Pathe E, Burley A. Posttraumatic stress disorder, secondary traumatic stress, and burnout in frontline workers in homelessness services: risk and protective factors. Journal of Social Distress and Homelessness. 2023 Mar 30;1–12. doi: 10.1080/10530789.2023.2191405

7. Wirth T, Mette J, Nienhaus A, Schillmöller Z, Harth V, Mache S. “This Isn’t Just about Things, It’s About People and Their Future”: a Qualitative Analysis of the Working Conditions and Strains of Social Workers in Refugee and Homeless Aid. International Journal of Environmental Research and Public Health. 2019 Oct 12;16(20):3858. doi: 10.3390/ijerph16203858

8. Hopper EK, Bassuk EL, Olivet J. Shelter from the Storm: Trauma-Informed Care in Homelessness Services Settings. The Open Health Services and Policy Journal. 2010 Apr 7;3(2):80–100. doi: 10.2174/1874924001003010080

9. Waegemakers Schiff J, Lane AM. PTSD Symptoms, Vicarious Traumatization, and Burnout in Front Line Workers in the Homeless Sector. Community Mental Health Journal. 2019 Jan 25;55(3):454–62. doi:10.1007/s10597-018-00364-7

10. Aykanian A, Mammah RO. Prevalence of Adverse Childhood Experiences Among Frontline Homeless Services Workers in Texas. Families in Society: The Journal of Contemporary Social Services. 2022 Feb 17;103(4):104438942110635. Doi: 10.1177/10443894211063579

11. van den Berk-Clark C. The Dilemmas of Frontline Staff Working With the Homeless: Housing First, Discretion, and the Task Environment. Housing Policy Debate. 2015 Apr;26(1):105–22. doi: 10.1080/10511482.2014.1003142

12. Olivet J, McGraw S, Grandin M, Bassuk E. Staffing Challenges and Strategies for Organizations Serving Individuals who have Experienced Chronic Homelessness. The Journal of Behavioral Health Services & Research. 2010 Jan 6;37(2):226–38. Doi: 10.1007/s11414-009-9201-3

13. Wirth T, Mette J, Prill J, Harth V, Nienhaus A. Working conditions, mental health and coping of staff in social work with refugees and homeless individuals: A scoping review. Health & Social Care in the Community. 2019 Mar 1;27(4):e257–69. doi: 10.1111/hsc.12730

14. Peters L, Hobson CW, Samuel V. A systematic review and meta-synthesis of qualitative studies that investigate the emotional experiences of staff working in homeless settings. Health & Social Care in the Community. 2021 Jul 13;30(1). doi: 10.1111/hsc.13502

15. Peter MD, Godfrey C, McInerney P, Munn Z, Tricco AC, Khalil H. Chapter 11: Scoping Reviews (2020 version). In: Aromataris E, Munn Z, editors. JBI Manual for Evidence Synthesis [Internet]. Joanna Briggs Institute; 2020 [cited 2024 Jul 29]. Available from: https://synthesismanual.jbi.global. doi: 10.46658/JBIMES-20-12

16. Arksey H, O’Malley L. Scoping studies: Towards a Methodological Framework. International Journal of Social Research Methodology. 2005;8(1):19–32. doi: 10.1080/1364557032000119616

17. Levac D, Colquhoun H, O’Brien KK. Scoping studies: Advancing the Methodology. Implementation Science. 2010 Sep 20;5(1):1–9. doi:10.1186/1748-5908-5-69

18. Tricco AC, Lillie E, Zarin W, O’Brien KK, Colquhoun H, Levac D, et al. PRISMA Extension for Scoping Reviews (PRISMA-ScR): Checklist and Explanation. Annals of Internal Medicine. 2018 Sep 4;169(7):467–73. 10.7326/m18-0850

19. Ng L. Well-being and burnout interventions for frontline homelessness staff: A Scoping Review Protocol [Internet]. OSF; 2023.Available from: osf.io/jp5yx

20. Hong QN, Pluye P, Fàbregues S, Bartlett G, Boardman F, Cargo M, et al. Mixed Methods Appraisal Tool (MMAT), Version 2018 (Registration of Copyright #1148552) [Internet]. Canadian Intellectual Property Office, Industry Canada; 2018 [cited 2024 Jul 29]. Available from: http://mixedmethodsappraisaltoolpublic.pbworks.com/w/file/fetch/127916259/MMAT_2018_criteria-manual_2018-08-01_ENG.pdf

21. Demasi RR. Helping the Helpers: Self-Care to Impact Compassion Fatigue in Volunteers Working With the Homeless [Internet] [Dissertation]. [Carlow University]; 2023 [cited 2024 Jul 29]. Available from: https://www.proquest.com/dissertations-theses/helping-helpers-self-care-impact-compassion/docview/2819153475/se-2?accountid=10673

22. Maguire N, Grellier B, Clayton K, Maguire N, Grellier B, Clayton K. The impact of CBT training and supervision on burnout, confidence and negative beliefs in a staff group working with homeless people. University of Southampton Institutional Repository [Internet]. 2017 [cited 2024 Jul 29]; Available from: https://eprints.soton.ac.uk/155113/

23. Munyoki N. Evaluation of a Mindfulness-Based Stress Reduction Program to Reduce Stress, Burnout, and Insomnia for Behavioral Healthcare Staff [Internet] [Dissertation]. [University of North Carolina]; 2022 [cited 2024 Jul 29]. Available from: https://www.proquest.com/dissertations-theses/evaluation-mindfulness-based-stress-reduction/docview/2671623364/se-2?accountid=10673

24. Jeffrey AC. Effect of feedback on levels of secondary traumatization of workers at battered women’s shelters across the United States [Internet] [Dissertation]. [Virginia Polytechnic Institute and State University]; 1999 [cited 2024 Jul 29]. Available from: https://vtechworks.lib.vt.edu/server/api/core/bitstreams/e6832175-bc04-4d0f-989b-0b362e2f2c38/content

25. Moore E, Soh M, Stuber M, Warde C. Well-being for trainees caring for homeless veterans. The Clinical Teacher. 2019 Jul 11;16(4). doi: 10.1111/tct.13053

26. Reeve A, Moghaddam N, Tickle A, Young D. A brief acceptance and commitment intervention for work-related stress and burnout amongst frontline homelessness staff: A single case experimental design series. Clinical Psychology & Psychotherapy. 2021 Mar 25;28(5). doi: 10.1002/cpp.2555

27. Skivington K, Matthews L, Sharon Anne Simpson, Craig P, Baird J, Blazeby J, et al. A new framework for developing and evaluating complex interventions: Update of Medical Research Council guidance. International Journal of Nursing Studies. 2024 Jun 1;154:104705–5. doi: 10.1016/j.ijnurstu.2024.104705

28. FEANTSA. ETHOS Typology on Homelessness and Housing Exclusion [Internet]. FEANTSA. 2017 [cited 2024 Jul 29]. Available from: https://www.feantsa.org/en/toolkit/2005/04/01/ethos-typology-on-homelessness-and-housing-exclusion

29. Lundy T, Crawford J. Health and Wellness Outcomes of Intimate Partner Violence Support Workers: A Narrative Review. Trauma, Violence, & Abuse. 2024 Feb 14;23(5). doi: 10.1177/15248380241231604

30. Godin K, Stapleton J, Kirkpatrick SI, Hanning RM, Leatherdale ST. Applying systematic review search methods to the grey literature: a case study examining guidelines for school-based breakfast programs in Canada. Systematic Reviews. 2015 Oct 22;4(1). doi: 10.1186/s13643-015-0125-0

31. Ward RJ, Greenway FT, Maguire N. “It Is Good to See the Person As a Whole Person and … Continue to Improve Our Psychologically Informed Working”: A Thematic Analysis of Clinical Psychology Trainee Placements in Homelessness Settings. Health expectations. 2024 Jun 1;27(3). doi: 10.1111/hex.14121

32. Kim J, Chesworth B, Franchino-Olsen H, Macy RJ. A scoping review of vicarious trauma interventions for service providers working with people who have experienced traumatic events. Trauma, Violence, & Abuse. 2022 Mar 9;23(5):152483802199131. doi: 10.1177/1524838021991310

33. Ferris LJ, Jetten J, Johnstone M, Girdham E, Parsell C, Walter ZC. The Florence Nightingale Effect: Organizational Identification Explains the Peculiar Link Between Others’ Suffering and Workplace Functioning in the Homelessness Sector. Frontiers in Psychology. 2016 Jan 28;27(3). doi: 10.3389/fpsyg.2016.00016

34. Hsu CC, Sandford B. The Delphi Technique: Making Sense of Consensus. Practical Assessment, Research, and Evaluation. 2007;12(1):10. doi: 10.7275/pdz9-th90

35. Cleland JA. The Qualitative Orientation in Medical Education Research. Korean Journal of Medical Education. 2017 May 29;29(2):61–71. doi: 10.3946/kjme.2017.53

36. Kubzansky LD, Kim ES, Boehm JK, Davidson RJ, Huffman JC, Loucks EB, et al. Interventions to Modify Psychological Well-Being: Progress, Promises, and an Agenda for Future Research. Affective Science. 2023 Mar 3;4(1). doi: 10.1007/s42761-022-00167-w

